# Uncovering the Genetic Architecture of Optic Nerve Integrity Estimates through Genome-wide Association Study Meta-analyses

**DOI:** 10.64898/2026.02.03.26345426

**Authors:** Asma M. Aman, Santiago Diaz-Torres, Samantha Sze-Yee Lee, Sjoerd J. Driessen, Victor A. de Vries, Frank C.T. van der Heide, Antonia Kolovos, Joshua M. Schmidt, Henry N. Marshall, Lania Saleh, Alicia Schulze, Gabriëlla AM. Blokland, Carroll A.B. Webers, Carla J.H. van der Kallen, Anke Wesselius, Ilja Arts, Freekje van Asten, Mathias Gorski, Martina E. Zimmermann, Klaus J. Stark, Iris M. Heid, Terri L. Young, Louis R. Pasquale, Ayellet V. Segrè, Janey L. Wiggs, Anthony P. Khawaja, Alex W. Hewitt, Alexander K. Schuster, Tos T.J.M. Berendschot, Alberta A.H.J. Thiadens, Karin A. van Garderen, Caroline C.W. Klaver, Pirro G. Hysi, Christopher J. Hammond, Caroline Brandl, Jamie E. Craig, Wishal D. Ramdas, Stuart MacGregor, David A. Mackey, Puya Gharahkhani

## Abstract

We conducted the first genome-wide association meta-analyses of global and sectoral peripapillary retinal nerve fibre layer (pRNFL) thickness and Bruch’s membrane opening-minimum rim width (BMO-MRW), the major optic nerve head structural and neurodegeneration biomarkers, including up to 25,942 and 12,080 participants, respectively, from the International Glaucoma Genetics Consortium. We identified 9 global pRNFL thickness and 9 global BMO-MRW loci, along with 28 and 19 loci for pRNFL and BMO-MRW sectors, respectively, comprising both shared and sector-specific loci. To identify intraocular pressure (IOP)-independent drug targets, global pRNFL thickness and BMO-MRW were conditioned on IOP. IOP-independent loci were then prioritised to identify candidate causal genes using transcriptome-wide association study and colocalization analysis. Several genes, such as *NMNAT2* and *TRIOBP,* had robust associations with both phenotypes, with potential IOP-independent therapeutic translation for glaucoma. Overall, we identified novel loci for pRNFL thickness and BMO-MRW, highlighting potential drug-target genes acting independently from IOP, and elucidating genetic differences among pRNFL sectors.

## Background

Glaucoma is a neurodegenerative eye disease that can lead to irreversible loss of vision and affects ∼3% of the world’s population^1^. It is characterised by progressive retinal ganglion cell (RGC) atrophy, often linked to increased intraocular pressure (IOP)^2^. Current glaucoma treatment focuses only on lowering IOP to prevent further damage, and does not reverse lost vision. Although IOP remains the major modifiable risk factor, many patients continue to experience progressive optic neuropathy despite optimal IOP control. As such, the development of novel neuroprotective interventions could provide new avenues to halt or slow RGC loss and preserve visual function.

RGC integrity estimates provide a framework to map the genetic architecture of glaucoma endophenotypes, offering deeper insight into the genetic drivers underlying optic nerve damage. Optical Coherence Tomography (OCT) allows precise measures of retinal and optic nerve structure. The peripapillary retinal nerve fibre layer (pRNFL) thickness and Bruch’s membrane opening-minimum rim width (BMO-MRW) are precise parameters for diagnosing glaucoma and monitoring progression, functioning as clinical biomarkers for assessing optic nerve health^3^. The pRNFL is the retinal layer surrounding the optic disc and contains the axons of the RGCs. Integrity is vital for transmitting visual information to the brain. In addition, BMO-MRW quantifies the minimum neuroretinal rim width measured from the BMO to the inner limiting membrane, providing insights into structural changes associated with glaucoma (**Figure 1**).

**Figure 1.**
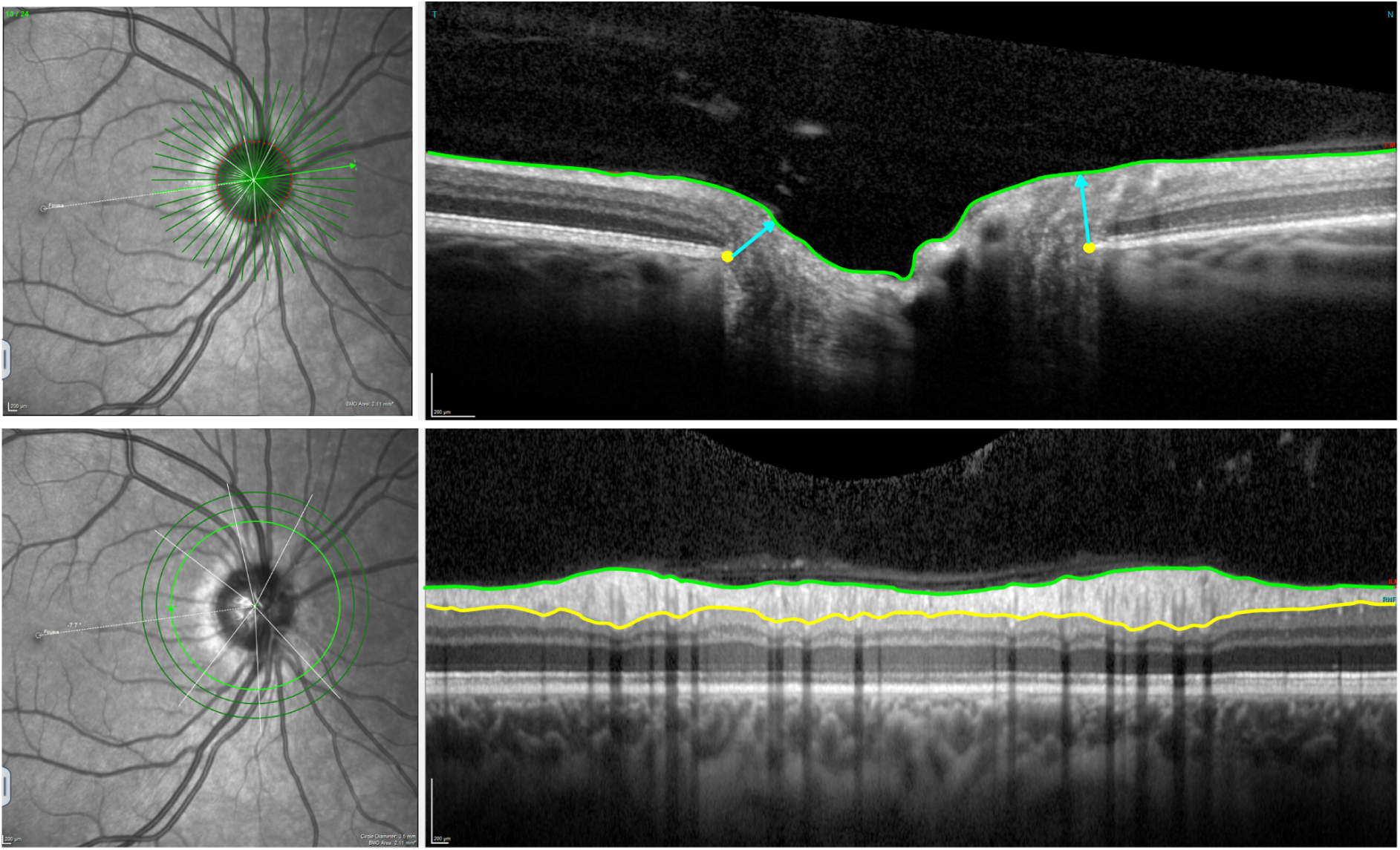
Example of a Heidelberg Spectral-Domain Optical Coherence Tomography radial-circle scan to obtain the BMO-MRW and pRNFL thickness. Top images: the BMO-MRW is indicated by the blue lines; 24 radial scans spaced 7.5° apart to quantify BMO-MRW, defined as the minimum distance between the Bruch’s membrane opening (yellow dots) and inner limiting membrane (green line). Bottom images: The pRNFL is delineated by the inner limiting membrane (green line) and the ganglion cell layer (yellow line). Circle scans provide pRNFL thickness measurement at the edge of a ∼3.5mm-diameter circle centred on the optic disc.

While significant progress has been made in genetic research on glaucoma, most loci identified to date are linked to IOP and vertical cup-to-disc ratio (VCDR)^4,5^, where the latter represents the currently explored neurodegenerative component of glaucoma. VCDR, a clinical parameter obtained from ophthalmoscopy and optic nerve photographs, is widely used to diagnose glaucoma and monitor its progression. BMO-MRW is a more precise measure of optic nerve health that could provide additional insight.

This study aimed to conduct the first genome-wide association study (GWAS) meta-analyses to identify genetic variants associated with pRNFL thickness and BMO-MRW. By combining OCT and genetic data from multiple cohorts within the International Glaucoma Genetics Consortium (IGGC), we sought to disentangle their genetic architecture and identify potential causal genes and pathways. The findings of this analysis could advance our understanding of the biology underlying both phenotypes which might possess implications in neurological maintenance and survival, thereby informing neuroprotective strategies for treating glaucoma.

## Results

### GWAS meta-analyses

#### Global pRNFL thickness and BMO-MRW

Nine loci were identified for each of the pRNFL thickness and BMO-MRW using 25,942 and 12,080 European participants, respectively (**Supplementary Tables 1-2**). Three of the genome-wide significant loci discovered for pRNFL thickness in this study have not been previously reported in association with macular RNFL^6^. Genetic correlation between pRNFL and macular RNFL thicknesses were only moderate (rg= 0.53, SE= 0.078, p-value= 2×10^-11^), indicating that a substantial genetic component of pRNFL is unique. The single nucleotide polymorphism (SNP) rs115803211 (near *TMEM161B*) in global pRNFL thickness was previously associated with glaucoma. The variant rs7916410 (near *ATOH7*) was associated with the optic disc area. In addition, the SNP rs1042602, which is mapped to the *TYR*, was reported in association with IOP and glaucoma. For global BMO-MRW, all loci have been associated with VCDR. The variants rs581876, rs8015152, rs1345467, and rs5750482 near *CDKN2B-AS1*, *SIX6*, *HNRNPA1L3*, and *TRIOBP*, respectively, were previously reported in association with glaucoma. Notably, adjustment for disc area had minimal impact on pRNFL, whereas BMO-MRW lost three loci after adjustment. The Manhattan plots are displayed in **Figure 2** for pRNFL thickness and BMO-MRW, and quantile-quantile (Q-Q) plots are presented in **Supplementary Figure 1**.

**Figure 2.**
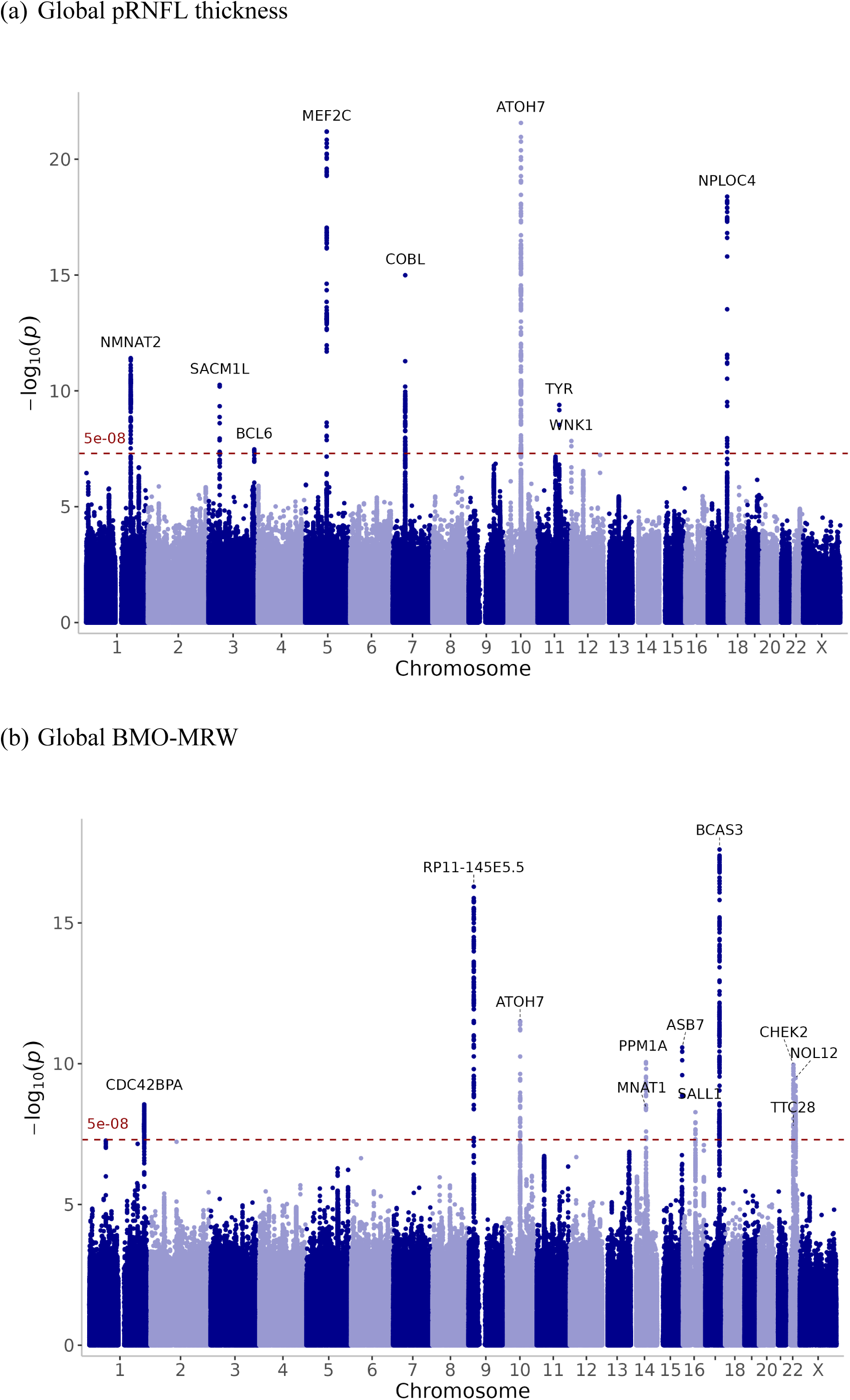
GWAS meta-analysis results for (a) global pRNFL thickness and (b) global BMO-MRW. Manhattan plot showing the –log₁₀ (p-values) of association tests in the Y-axis. Each point represents an SNP plotted by chromosomal position. The horizontal dashed red line indicates the genome-wide significance threshold (p-value = 5 × 10⁻⁸). The nearest gene was annotated for the top SNPs.

#### Sectoral pRNFL and BMO-MRW

Global pRNFL thickness and BMO-MRW are the weighted average across six sectors (nasal, temporal, superonasal, superotemporal, inferonasal, and inferotemporal). The analysis of pRNFL sectors in 19,763 participants revealed in total 28 independent genomic risk loci, including shared and sector-specific associations, as illustrated in the circular Manhattan plot (**Figure 3**). Notably, four pRNFL sectoral regions (temporal, superotemporal, inferonasal, and inferotemporal) had a genome-wide significant locus on chromosome 14, near *SIX6* gene, which is a well-known optic nerve gene, that was not found in the global pRNFL. We observed that the effect sizes of the top SNP at this locus had opposite directions between the temporal and nasal regions (**Supplementary Table 3**). As a result, this cancelled out their effects in the global pRNFL GWAS. In addition, there was a significant difference in the effect sizes between the temporal region and other pRNFL sectors for rs17421627. This SNP is located at the locus near *TMEM161B/MEF2C* genes on chromosome 5, which is commonly associated with retinal vasculature measurements^7^. Conversely, other signals (e.g., rs77674430 and rs72957725) were unique to the temporal region and showed significantly different effect sizes compared to other sectors, as presented in **Supplementary Table 3**.

**Figure 3.**
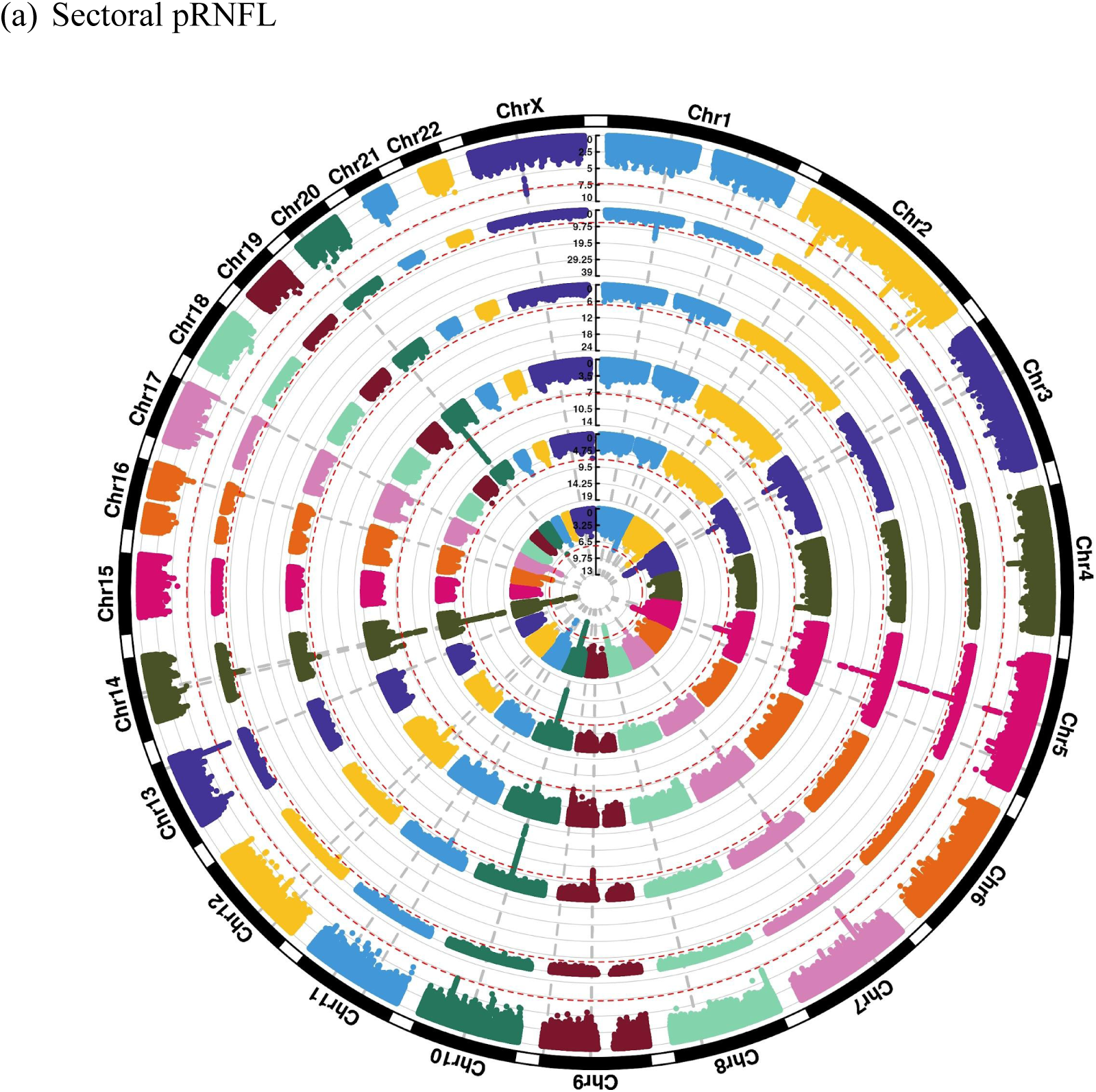
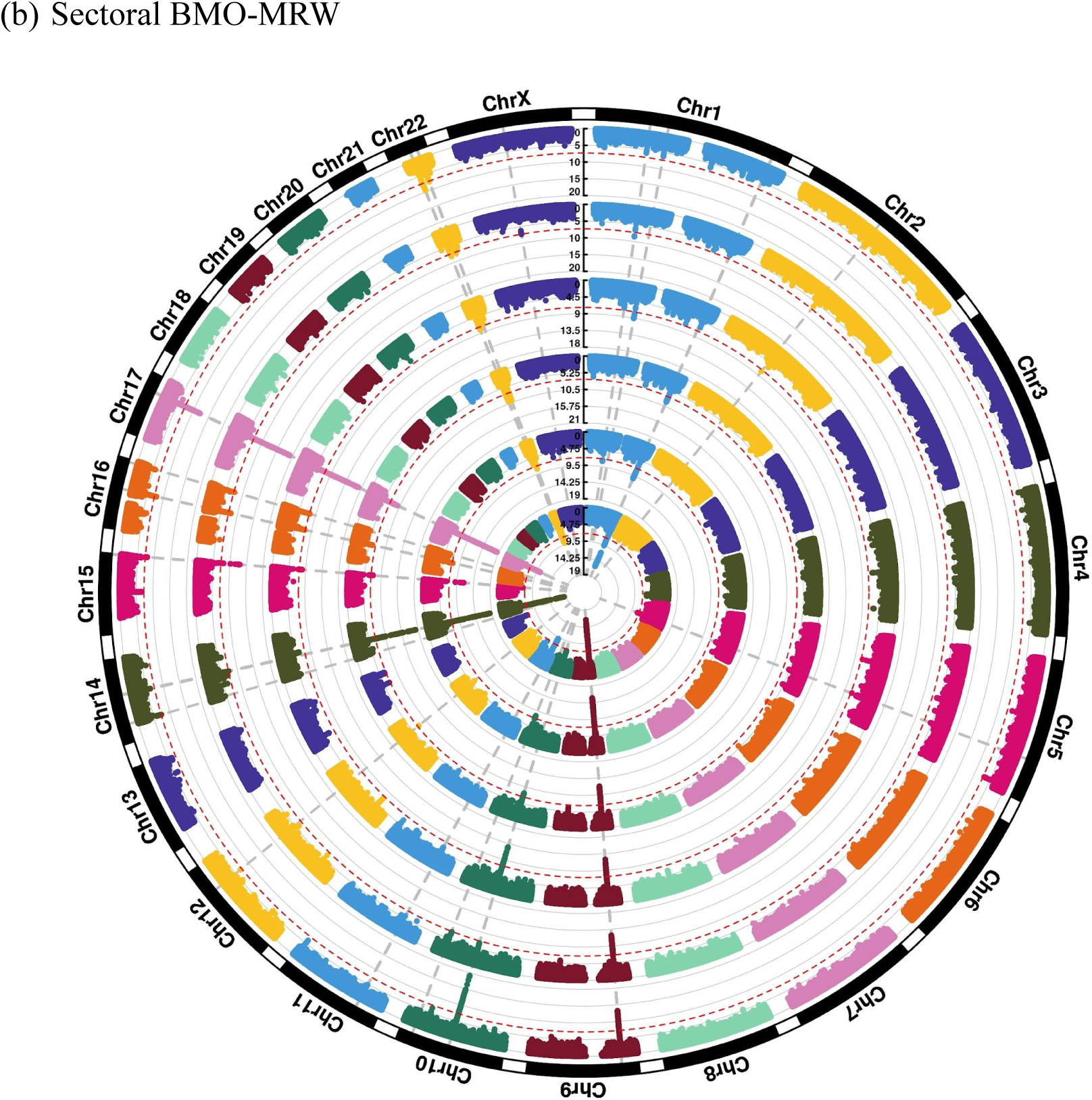
Circular Manhattan plots showing (a) sectoral pRNFL and (b) sectoral BMO-MRW. The order of sectors, starting from the inner circle, is as follows: temporal, inferotemporal, superotemporal, nasal, inferonasal, and superonasal. The grey vertical lines refer to genomic risk loci, while the red circular lines refer to genome-wide significance threshold (p-value = 5 × 10⁻⁸).

From the analysis of BMO-MRW sectors in 12,080 individuals, a total of 19 genomic risk loci were identified, comprising shared and distinct loci, as shown in the circular Manhattan plot (**Figure 3**). Interestingly, the temporal regions’ effect sizes near *SIX6* were roughly triple the effect sizes of those in the nasal regions but in the same direction of effect (**Supplementary Table 4**). In addition, the variants rs3118379 (near *RPE65*) and rs3125620 (near *HSPA12A*) were previously reported for glaucoma. The SNP rs3118379 exhibited genome-wide significance exclusively in the inferotemporal and inferonasal regions, whereas rs3125620 reached genome-wide significance level only in the inferotemporal sector. Manhattan plots for pRNFL and BMO-MRW sectoral regions are provided in the **Supplementary Figure 2**. Genetic correlation results across sectors are presented in a later section. These results suggest that although some risk loci have similar effects across sectors, many loci display sector-specific differences in effect size or act uniquely within a given sector.

#### BMO area

Using data from 12,068 participants, we identified 11 loci for BMO area (i.e., the area within the anatomical border of the optic nerve head), all of which have previously been associated with optic disc area^8^. The Manhattan plot is presented in the **Supplementary** Figure 2.

#### Gene-based and Pathway-based analyses

The mBAT-combo analysis mapped 19 genes within seven distinct loci to global pRNFL (**Supplementary Table 5**), where the strongest associations were observed on chromosome 10, including genes such as *MYPN*, *ATOH7*, *PBLD,* and *HNRNPH3*. Additionally, a locus in chromosome five was implicated and mapped to *TMEM161B*.

For global BMO-MRW, a total of 20 genes within eight distinct loci were mapped by mBAT-combo, as detailed in the **Supplementary Table 6.** *OR4C16* and *OR4C11* located on chromosome 11 were the most significant genes identified. In addition, the genes *CDKN2B*, *CDKN2A*, and *SIX6* were mapped to BMO-MRW and were previously associated with overall glaucoma^5,9,10^ and normal tension glaucoma^11^. None of the pathways survived multiple testing corrections in the MAGMA pathway analysis conducted for pRNFL thickness and BMO-MRW.

#### Potential neurodegenerative loci independent from IOP

Using the multi-trait-based conditional and joint association analysis (mtCOJO) method, nearly all global pRNFL thickness loci retained their associations after conditioning on VCDR and IOP. Similarly, global BMO-MRW loci were mostly independent of IOP; however, they were entirely overlapping with VCDR. Manhattan plots for IOP-adjusted pRNFL thickness and BMO-MRW are shown in **Supplementary** Figure 3. Transcriptome-wide association study (TWAS) analysis revealed, using blood and retinal tissues, 76 genes for IOP-adjusted pRNFL thickness, while 23 genes were identified for IOP-adjusted BMO-MRW. Based on subsequent colocalization analysis, variants affecting *MRPL48*, *TUBA1C,* and *NMNAT2* gene expression and IOP-adjusted pRNFL thickness were colocalized, each with a posterior probability > 50% (**Table 1**). Similarly, the colocalization analysis prioritised several genes for IOP-adjusted global BMO-MRW, including *PBLD*, *TRIOBP*, *H1F0,* and *HAUS4* (**Table 1**), suggesting robust associations with BMO-MRW. The full list of genes identified through TWAS are available in **Supplementary Tables 7-10**.

**Table 1.**
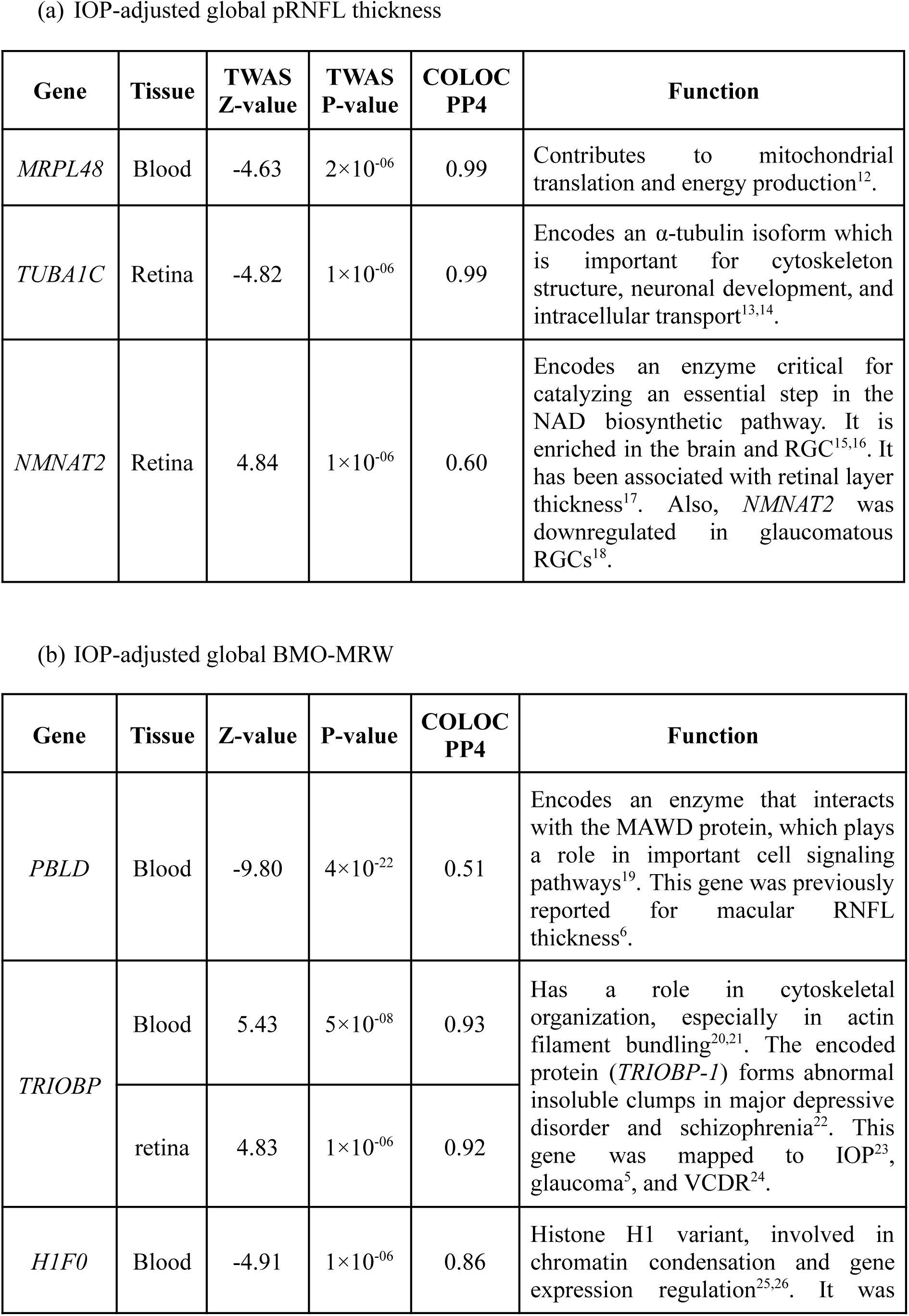
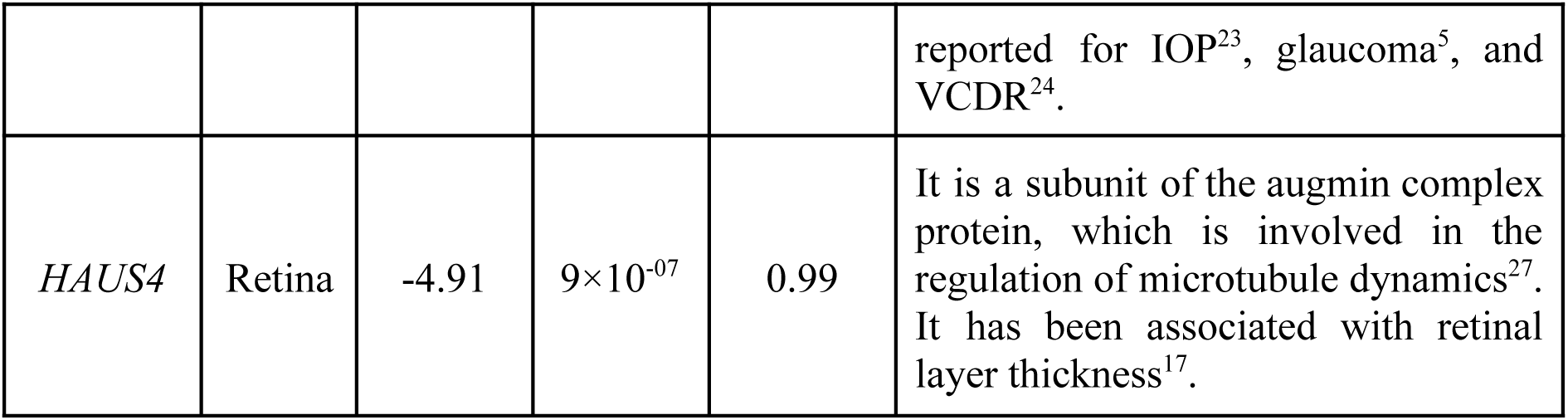
Overview of genes prioritised by TWAS and colocalization.

#### Genetic correlation

The LD score regression (LDSC) analysis revealed a moderate inverse genetic correlation between glaucoma and global pRNFL thickness (rg= −0.237, SE= 0.048, p-value= 6×10^-07^). Furthermore, the genetic correlation between glaucoma and the various pRNFL sectoral regions was comparable to that of global pRNFL (**Supplementary Table 11**), except for the nasal region (rg= −0.06, SE= 0.049, p-value= 0.22), which showed a nominally significant difference in its correlation with glaucoma compared to global pRNFL (difference z-score=-2.607, p-value= 0.009). The differential strengths of genetic correlation between glaucoma and the various pRNFL sectors is consistent with clinical expectations, where the nasal sector is one of the last to be affected in glaucoma. The pairwise genetic correlations revealed strong associations among temporal regions (rg ranging between 0.59 and 0.73) and moderate associations within nasal regions (rg ranging between 0.33 and 0.58). Temporal sectors showed weak or negligible genetic correlations with the nasal sectors (**Supplementary Table 12**).

Global BMO-MRW showed strong genetic correlation with glaucoma (rg= −0.689, SE= 0.051, p-value= 9×10^-42^), with consistent genetic correlation estimates with glaucoma across sectoral BMO-MRW measurements (**Supplementary Table 13**), indicating that the genetic components for sectoral BMO-MRW are similar. All six sectors of BMO-MRW also showed very strong and highly significant genetic correlation between each other (**Supplementary Table 14**).

Furthermore, global pRNFL and global BMO-MRW were weakly associated (rg= 0.104, SE= 0.093, p-value= 0.26). Global pRNFL showed significant moderate genetic correlation with VCDR (rg= −0.231, SE= 0.062, p-value= 2×10^-04^), whereas the correlation with IOP did not survive multiple testing (rg= −0.153, SE= 0.050, p-value= 0.002). In addition, there was a very strong genetic correlation between global BMO-MRW and VCDR (rg= −0.981, SE= 0.048, p-value= 1×10^-92^) and a moderate correlation with IOP (rg= −0.207, SE= 0.051, p-value= 5×10^-05^).

Regarding the BMO area, there was a moderate genetic correlation with VCDR (rg= 0.299, SE= 0.060, p-value= 8×10^-07^) and a weak but nominally significant genetic correlation with glaucoma (rg= 0.111, SE= 0.049, p-value= 0.02). In addition, the correlation was weak and not statistically significant between BMO area and refractive error (rg= 0.062, SE= 0.037, p-value= 0.10).

#### Causal inference

Using the inverse variance weighted (IVW) method, we observed significant causal effects of IOP on pRNFL thickness (effect size= −0.037, SE= 0.009, p-value= 3×10^-05^), and on BMO-MRW (effect size= −0.130, SE= 0.014, p-value= 9×10^-22^). Also, Mendelian randomisation (MR) sensitivity analysis supported the IVW results, where the effect sizes remained consistent across all approaches, supporting the robustness of the findings (**Figure 4**; **Supplementary** Figure 4). Notably, the IOP causal effect size on BMO-MRW was significantly greater than the effect on pRNFL thickness (z-score= 5.67, p-value= 1×10^-08^).

**Figure 4.**
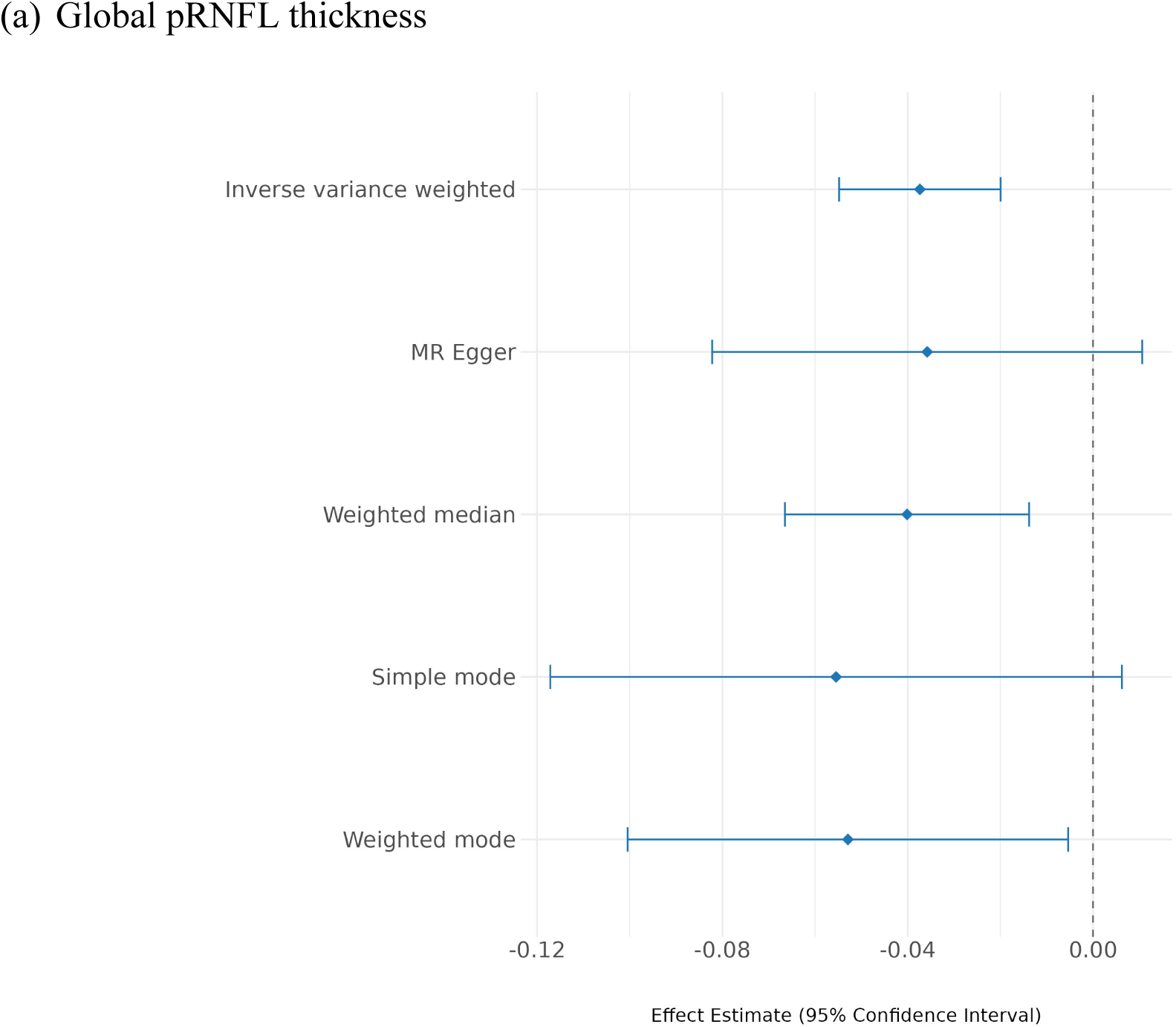
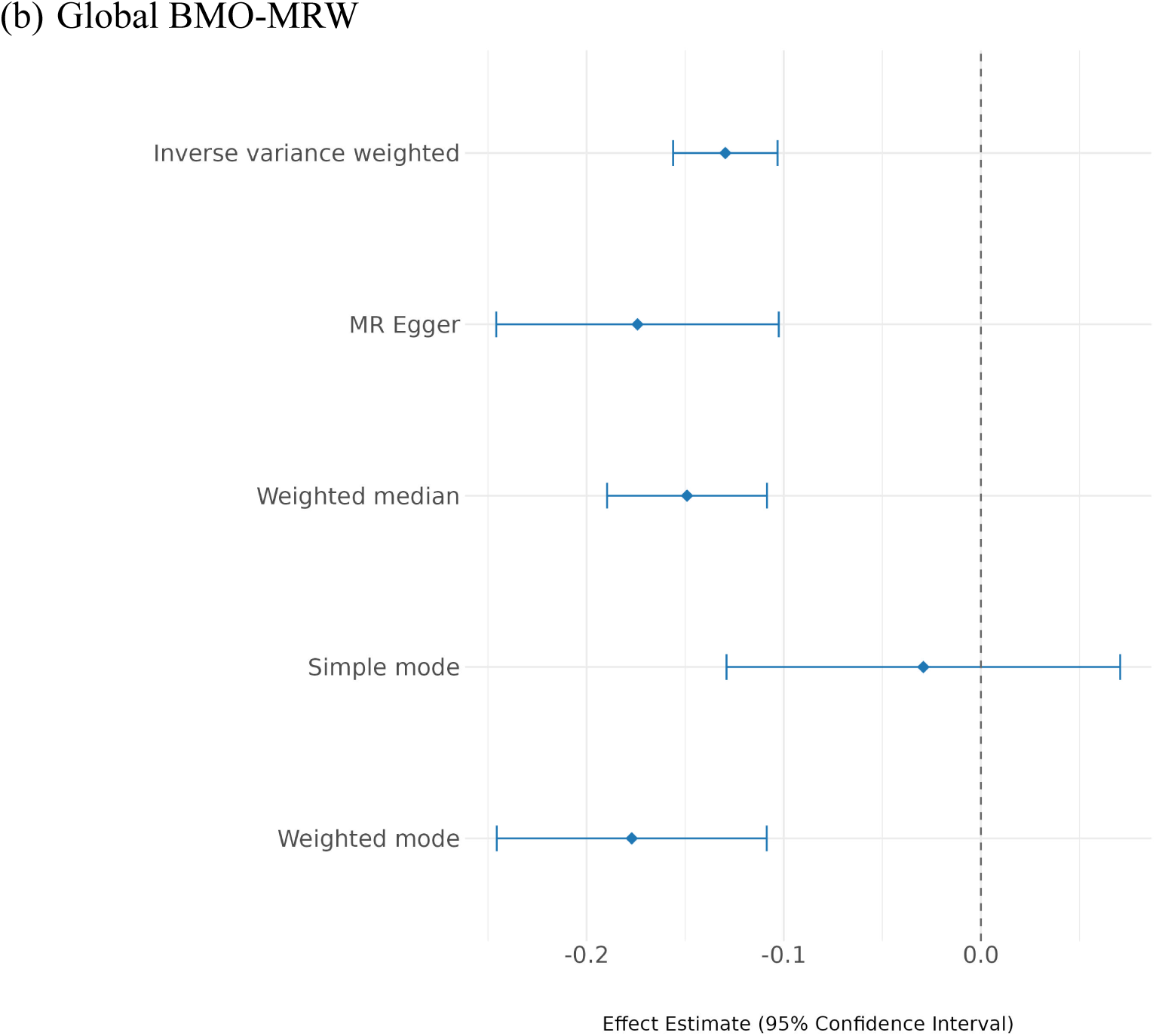
Mendelian randomisation results among different methods for IOP on:

## Discussion

The pRNFL thickness and BMO-MRW are established biomarkers for diagnosing glaucoma and monitoring disease progression. This study lays the foundation for elucidating the genetic basis of global and sectoral pRNFL thickness and BMO-MRW. By combining data from different cohorts, we identified several loci for glaucoma endophenotypes, allowing us to further shed light on the disease’s neurodegenerative aetiology. Moreover, valuable insights into the genetic differences between retinal sectors were revealed.

Our MR analyses support a causal effect of IOP on both pRNFL and BMO-MRW, consistent with previous studies reporting that higher IOP is associated with pRNFL and BMO-MRW thinning^28–30^. Notably, the estimated causal effect on BMO-MRW was substantially greater than that on pRNFL, suggesting that BMO-MRW may be more sensitive to changes in IOP. This is further supported by a study that investigated the responsiveness of BMO-MRW to acute IOP elevation^30^. From another perspective, mtCOJO conditioning showed that most genetic loci associated with pRNFL thickness and BMO-MRW remain significant after adjusting for IOP, suggesting that these traits related loci are not IOP-driven rather they act directly on the nerve head^11^. This is also supported by a modest negative genetic correlation with IOP, indicating partial but not strong genetic overlap. Genetic independence between IOP and pRNFL thickness, as well as BMO-MRW, suggests that glaucoma susceptibility involves both mechanical damage from elevated IOP and intrinsic vulnerability of the optic nerve. Therefore, pRNFL thickness and BMO-MRW may represent neurodegenerative aspects of glaucoma, which can be targeted for developing neuroprotective drugs.

From the genetic correlation findings, we observe that BMO-MRW and VCDR share substantial genetic influences on optic nerve head structure. Although VCDR is a widely used clinical parameter for glaucoma diagnosis, it is subjective and depends on the clinical assessment and the experience level of the healthcare professional rather than on clear objective standards, putting it at risk of inter-observer variability^31,32^. Furthermore, AI-predicted VCDR is more accurate than manually graded values^4^. In contrast, BMO-MRW is an objective and reproducible quantification of neuroretinal rim tissue thickness, providing precise structural information about the optic nerve head. It is also a sensitive biomarker for detecting early glaucomatous changes^30^.

Previous research has demonstrated anatomical and histological differences in pRNFL thickness between the various sectors. In healthy eyes, the pRNFL is typically thicker in the inferior and superior sectors due to the concentration of ganglion cell axons in the arcuate fiber areas^33^. These sectoral differences are clinically relevant, as the thinning patterns serve as early signs for some optic neuropathies. For instance, inferior and superior regions thinning are commonly observed in early glaucomatous optic neuropathy^34–36^. Our findings align with these observations, where we observed differences among pRNFL sectors with their correlation with each other and with glaucoma. In addition, the nasal temporal difference associated with *SIX6* was previously noted in 1,598 members of the TwinsUK study (mean age 63 years) with a thicker nasal sector and thinner temporal sector observed with SNPs rs1043727 and rs33912345^37^. A similar analysis of 943 members of the Raine Study Gen2 (mean age 20 years) found no associations^37^. *SIX6* is also associated with myopia^38^, a well-known risk factor for glaucoma. Whether myopia induces tilting of the optic nerve to create this asymmetric pRNFL change warrants further investigation.

TWAS and colocalization analyses support the association between pRNFL thickness and a few genes, including *NMNAT2*. This gene encodes an enzyme responsible for converting NMN (nicotinamide mononucleotide) to NAD⁺ ^15^, where the latter plays a crucial role in neuronal health by protecting the cells from oxidative stress. In single cell analyses, *NMNAT2* was highly expressed in retinal neuronal cells, particularly in RGCs. The expression differed among individual samples, with lower expression correlated with increased susceptibility to neurodegeneration in mouse models of glaucoma^39^. Nicotinamide levels were observed to be lower in glaucoma patients compared to controls^40^. Furthermore, NAD levels in mice declined with age, which increased the vulnerability of RGCs to degeneration due to stressors such as elevated IOP^41^, while *NMNAT2* levels influenced RGC susceptibility to neurodegeneration^39^. *NMNAT2* was presented as a target for neuroprotection, given that delivering human *NMNAT2* via gene therapy conferred a strong neuroprotection to RGC in mice^39^, and overexpression of this gene enhanced RGCs survival and protected vision in glaucoma eyes^18^. Similarly, previous research found that increasing the enzyme levels may have neuroprotective effects on glaucoma^42,43^. Administering nicotinamide and pyruvate supplements for glaucoma treatment has already reached a phase ll clinical trial^44^, supporting its potential role in neuroprotection. A recent GWAS of macular thickness in the UK Biobank identified an association with two SNPs within *NMNAT2*; rs12035399 and rs2788045^17^. Testing participants in NAD clinical trials for these *NMNAT2* variants may be useful in identifying levels of response to NAD treatment.

TWAS and colocalization also highlighted a few genes, e.g., *TRIOBP*, associated with BMO-MRW. This gene is likely a pleiotropic gene that has effects on both IOP and BMO-MRW. In particular, a previous study demonstrated that knockout of *TRIOBP* gene was associated with elevated levels of IOP^45^. In addition, our TWAS analysis showed that *TRIOBP* expression was associated with thicker BMO-MRW mostly independently of the IOP pathway, as shown in the mtCOJO results. These observations are supported by a recent study, which showed that the expression of *TRIOBP* has a protective effect on POAG^46^. This gene holds a promising future as a drug target for neuroprotection.

This study reports novel GWAS hits for pRNFL thickness and BMO-MRW. However, it has a few limitations. For instance, the GWAS was restricted to individuals of European ancestry, which limits the generalizability of the findings to other populations. Future GWAS should include participants from other ancestries to validate the findings across diverse populations and to reveal potential ancestry-specific genetic associations. In addition, the sample size of sectoral pRNFL thickness was small compared to the global phenotype. As a result, the statistical power to detect weak signals was reduced. Similarly, the limited sample size of the EyeGEx dataset may have reduced the power to discover additional genes in TWAS. Furthermore, functional analysis should be conducted to identify potential neuroprotective treatment targets for glaucoma.

In conclusion, we identified novel loci for glaucoma-related neurological traits that could serve as a base for future investigations to discover neuroprotective drug targets. Furthermore, we elucidated the genetic differences among pRNFL sectors with potential relevance to clinical practice.

## Methods

### Study cohorts

The IGGC is a global initiative focused on uncovering glaucoma genetic risk factors across diverse populations. We included 10 cohorts for global pRNFL GWAS meta-analysis and five cohorts for global BMO-MRW GWAS meta-analysis to identify associated genetic variants. GWAS analyses were conducted for each cohort according to a standardised analysis plan, as described below, to ensure consistency and minimise heterogeneity. The total sample size was 25,942 and 12,080 participants for global pRNFL and global BMO-MRW, respectively. All included participants were of European descent. For sectoral GWASs, data were available from 19,763 and 12,080 individuals for pRNFL and BMO-MRW, respectively. In addition, 12,068 participants were included for the BMO area. Further details about study-specific characteristics can be found in the **Supplementary Tables 15-18**.

### Phenotype measurement

The pRNFL thickness and BMO-MRW were measured using OCT scans. The OCT devices capture multiple cross-sectional images of the retina using light waves to measure sectoral thickness. The optic disc was analysed in six standard sectors: nasal, temporal, superonasal, superotemporal, inferonasal, and inferotemporal, as illustrated in **Figure 5**. The weighted average value across all sectors was calculated by the OCT analysis software used in each study to obtain the global pRNFL thickness and global BMO-MRW. The specific OCT model and protocol employed by each study is detailed in the **Supplementary Table 19**.

**Figure 5.**
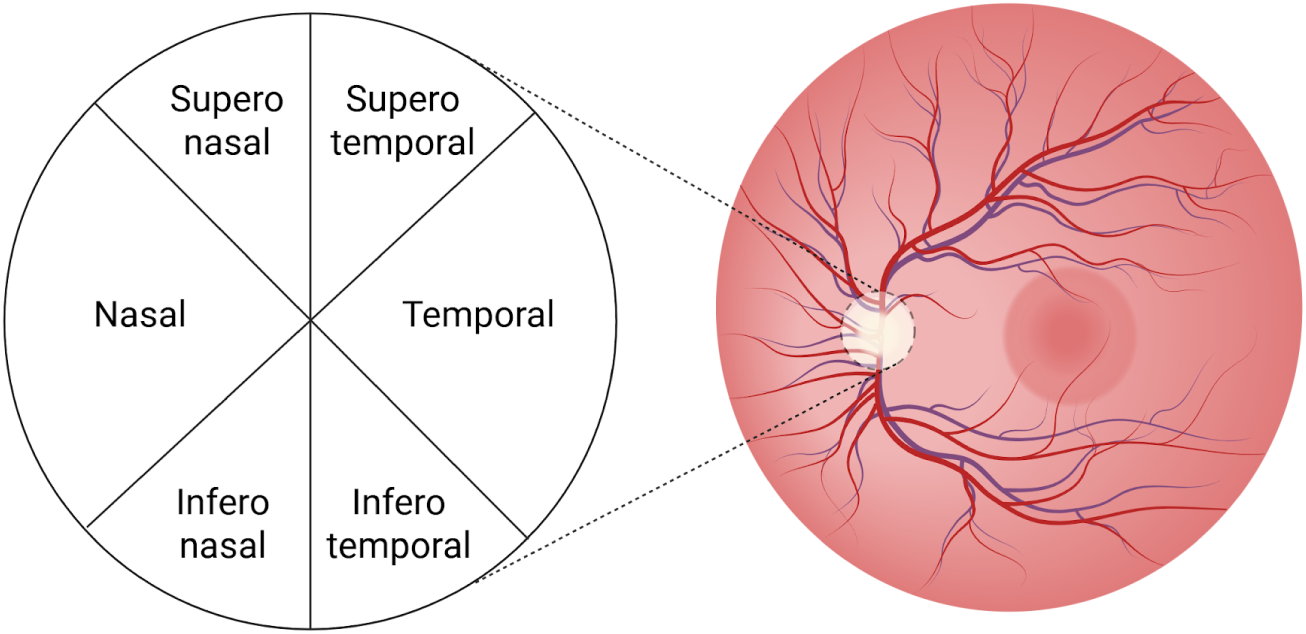
Retinal image of the left eye showing the optic disc divided into six sectors (nasal, temporal, superonasal, superotemporal, inferonasal, and inferotemporal).

In this study, the primary phenotypes were the global pRNFL thickness and global BMO-MRW, while the secondary phenotypes consisted of BMO area, sectoral pRNFL thickness, and sectoral BMO-MRW. For each participant, the mean global values from both the right and left eyes were calculated when data from both eyes were available. Each study applied a rank-based inverse normal transformation to the measurements to minimise the potential heterogeneity when combining the results in the meta-analysis.

### Genotyping and quality control

Details about genotyping and imputation methods carried out by each study can be found in the **Supplementary Table 19**. All datasets underwent similar quality control criteria, including filtering out individuals with more than 0.02 missingness of genotype data, relatedness pi-hat more than 0.1, and ancestral outliers whose first or second principal components were more than six standard deviations from the European mean, with reference to the 1000 Genomes European centroid. Similarly, genotypes were filtered based on the following criteria: Hardy-Weinberg Equilibrium p-value less than 10^-6^, missingness rate exceeding 0.05, and minimum allele frequency (MAF) below 0.01.

### GWAS

All studies used linear regression models to conduct GWAS for the transformed phenotypes. The model conducted association tests for each genetic variant while adjusting for covariates including age, sex, the first 10 principal components, OCT model if more than one was used in the same study, and axial length (i.e., the distance from the cornea to the retina) or spherical equivalent (i.e., a value that summarizes the eye’s overall focusing power) if axial length was not available. The latter covariate accounted for transverse magnification during the OCT image process due to variations in the length of the eye.

### GWAS meta-analysis

We employed fixed effect models to conduct primary and secondary phenotypes GWAS meta-analyses using METAL software 02-11-2021 release^47^, which assumes that the effect of each SNP is consistent across different studies. GWAS quality control criteria included filtering genetic variants with MAF below 0.01 and an imputation quality score less than 0.3, ensuring that only high-quality and common SNPs were retained for further analysis. In addition, the quality check included allele frequency harmonisation with the UK Biobank reference panel, comprising 5,000 healthy (i.e., no records of ICD-10 disorders) and unrelated participants, to ensure consistency across datasets. Visualisation using Manhattan and Q-Q plots were utilised to assess the overall distribution of observed p-values for each SNP and detect potential biases.

We utilised the FUMA platform^48^ to identify independent genomic risk loci. They were defined based on a set of criteria including: genome-wide significance level (i.e., p-value below 5×10^-8^), SNP present within 1 Mb genomic distance, and clumping threshold r^2^ less than 0.2 using the European UK Biobank LD Reference panel. We used topr^49^ and CMplot^50^ packages in R to create annotated Manhattan plots and circular Manhattan plots, respectively. Also, BMO-MRW and pRNFL thickness were adjusted for optic disc area using the mtCOJO method^51^ to estimate SNP effects independent from disc size.

### Gene-based association analysis

Gene-based association analyses were conducted using mBAT-combo^52,53^, a method that combines test statistics across multiple genetic variants within each gene to assess their collective association with the phenotype of interest. It has the potential to identify genes that are usually missed because of SNPs with masking effects. We applied mBAT-combo to the GWAS summary statistics to identify genes significantly associated with the primary phenotypes, adjusting the significance threshold for multiple testing using the Bonferroni method (p-value < 0.05/ (18,992 genes × 2 traits tested)). The LD reference panel consisted of ∼5,000 healthy participants from the UK Biobank.

### Pathway-based analysis

Pathway enrichment analyses were conducted using MAGMA^54^, which was implemented within the FUMA platform^48^ to identify biologically relevant pathways associated with global pRNFL thickness and global BMO-MRW. Curated gene sets from the Molecular Signatures Database (MsigDB v2023.1Hs) and gene ontology terms were used as the predefined gene set database. A Bonferroni correction was applied to correct for multiple testing based on the number of prespecified gene sets and number of traits tested (p-value < 0.05/ (17,023 gene sets × 2 traits tested)).

### Causal genes prioritisation

We employed the mtCOJO^51^ method to identify loci associated with global pRNFL thickness and global BMO-MRW that are independent of the effects of VCDR and IOP. Using GWAS summary data, we performed conditional GWAS analyses of global pRNFL thickness and global BMO-MRW, conditioning separately on VCDR^4^ and IOP^23^. Afterwards, we conducted TWAS on IOP-adjusted global pRNFL thickness and global BMO-MRW, obtained from mtCOJO results, to find genes independent of IOP pathways in order to identify neurodegenerative loci. TWAS is a method that integrates genetic and transcriptomic data to identify genes whose predicted expression levels are associated with complex traits. We utilised SUMMIT tool^55^, by integrating GWAS summary statistics for IOP-adjusted global pRNFL and IOP-adjusted global BMO-MRW with whole blood expression quantitative trait loci (eQTL) reference data obtained from eQTLGen consortium^56^. Similarly, we performed TWAS on retinal tissue eQTL data obtained from EyeGEx^57^ using FUSION^58^. We corrected the p-value threshold for multiple comparisons based on the number of genes in each tissue and the number of traits tested: p-value < 2.19×10^-6^ (i.e., 0.05/(11,433×2)) for whole blood and p-value < 1.46×10^-6^ (i.e., 0.05/(17,143×2)) for retina. The most likely causal genes were then prioritised using colocalization analysis via the coloc tool^59^. It estimates the posterior probability of five hypotheses whether gene expression and a trait (pRNFL thickness or BMO-MRW) share causal variants at a locus: H0) no association between gene expression and the trait; H1) the causal variant is associated with the gene expression only; H2) the causal variant is associated with the trait only; H3) distinct causal variants are associated with the gene expression and the trait; and H4) shared causal variant is associated with the gene expression and the trait. We reported genes passing the posterior probability threshold of 0.5.

### Genetic correlation

The genetic correlations were estimated using the bivariate LDSC method^60,61^, which was computed based on GWAS summary statistics and LD scores obtained from the UK Biobank reference panel. To identify the extent of genetic overlap between macular and peripapillary retinal nerve thickness layers, we estimated the genetic correlation between our global pRNFL and macular RNFL (N = 31,434)^6^. Furthermore, the genetic correlations were estimated between glaucoma, VCDR, IOP, global pRNFL thickness, and global BMO-MRW. For glaucoma, summary statistics were obtained from the multi-trait analysis of GWAS conducted for more than 600,000 individuals of European ancestry^5^. VCDR summary statistics were acquired from a GWAS conducted on AI-based gradings of VCDR values adjusted for vertical disc diameter from 111,724 participants^4^, while those for IOP were obtained from a GWAS meta-analysis involving 133,492 individuals^23^. In addition, genetic correlations were evaluated between glaucoma and sectoral pRNFL, as well as sectoral BMO-MRW. To explore sectoral differences, we compared the genetic composition among sectors by conducting pairwise genetic correlations. We also investigated the genetic correlation of the BMO area with glaucoma^5^, VCDR^4^, and refractive error. Summary statistics for refractive error were obtained from a GWAS meta-analysis with more than half a million European participants^62^. The significance level was adjusted for multiple testing using the Bonferroni method, applying a threshold of p-value < 0.0009 (i.e., 0.05/53), where 53 is the number of genetic correlation tests conducted across all trait pairs.

### Causal inference

We used two-sample MR to test causality of IOP^23^ (i.e., exposure) on global pRNFL thickness and BMO-MRW (i.e., outcomes). We used clumping to select instrumental variables for IOP using PLINK version 1.9, where we selected independent genome-wide significant SNPs by setting r^2^ to 0.001 and p1 to 5×10^-8^. The IVW method^63^ was used as the primary approach for the analysis. To assess the robustness of the findings, we conducted MR sensitivity analysis using Egger regression^64^, weighted median^65^, and both simple and weighted mode estimators^66^. To evaluate heterogeneity, we first computed the overall Q statistics. When significant heterogeneity was detected, we calculated Q statistics at the SNP level (χ², df = 1) and excluded SNPs that showed high heterogeneity from the MR analysis^67^. The TwoSampleMR library was used in R to conduct the analysis^68^. The Bonferroni method was applied to correct for multiple comparisons (p-value < 0.05/2, to account for two MR tests).

## Ethics

All contributing studies received ethical approval from their respective institutional review boards, and informed consent was obtained from all participants.

## Data availability

Data are available from The Maastricht, The AugUR, and ANZRAG Studies for any researcher who meets the criteria for access to confidential data; the corresponding author may be contacted to request data. Data described in the manuscript will be made available upon request pending (e.g., application and approval, payment, other). TwinsUK welcomes data sharing with health researchers via a managed data access process which can be accessed at https://twinsuk.ac.uk/researchers/access-data-and-samples/request-access/.

## Code availability

The code and AI model for feature extraction used in the Rotterdam Study is available with open source license: https://github.com/Eyened/OCT-ONH. No other custom code was generated for this study. All methods and tools are publicly available and appropriately referenced.

## Acknowledgement

The authors would like to acknowledge the Busselton Health Study participants, and the Raine Study participants and their families for their ongoing participation in the study. We thank the Raine Study team for study co-ordination and data collection, as well as the NHMRC and the Raine Medical Research Foundation for their long-term contribution to funding the Raine Study over the last 30 years. Also, the authors would like to acknowledge the ZIO foundation (Vereniging Regionale HuisartsenZorg Heuvelland) for their contribution to The Maastricht Study. The researchers are indebted to all participants for their willingness to participate in the study.

Figure 5 was created in BioRender. Diaz, S. (2025) https://BioRender.com/h0amnam

