## Supplementary Figures for "Uncovering the Genetic Architecture of Optic Nerve Integrity Estimates through Genome-wide Association Study Meta-analyses"

**Supplementary Figure 1.** Q-Q plot comparing the observed (x-axis) versus expected $-\log_{10}$(p-values) (y-axis) under the null hypothesis of no association.

(a) Global pRNFL thickness

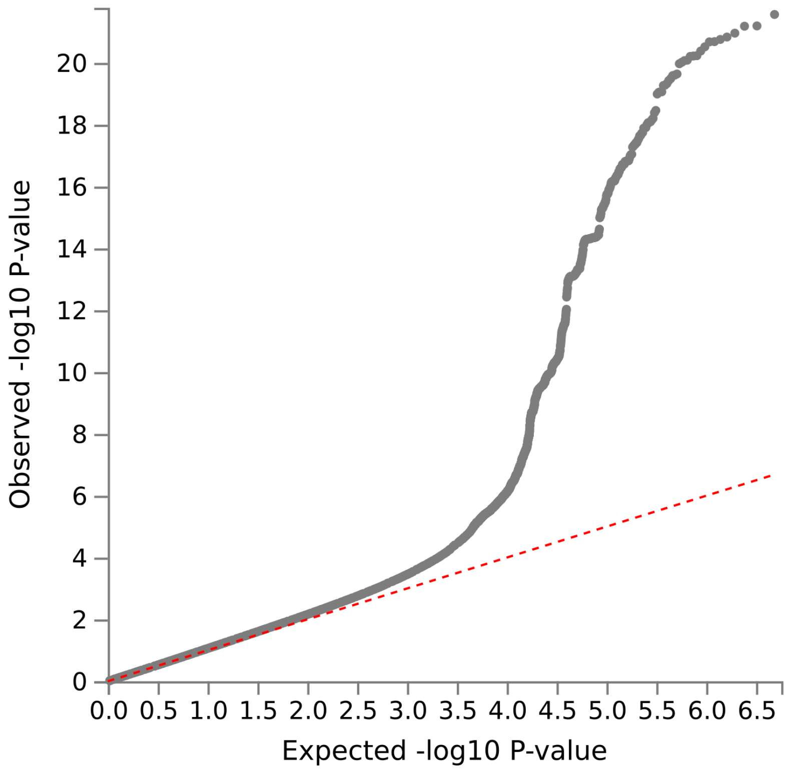


- Lambda GC: 1.0957
- Mean $Chi^2$: 1.12
- Intercept: 1.0304 (0.0075)
- Attenuation ratio: 0.2536 (0.0622)

(b) Global BMO-MRW

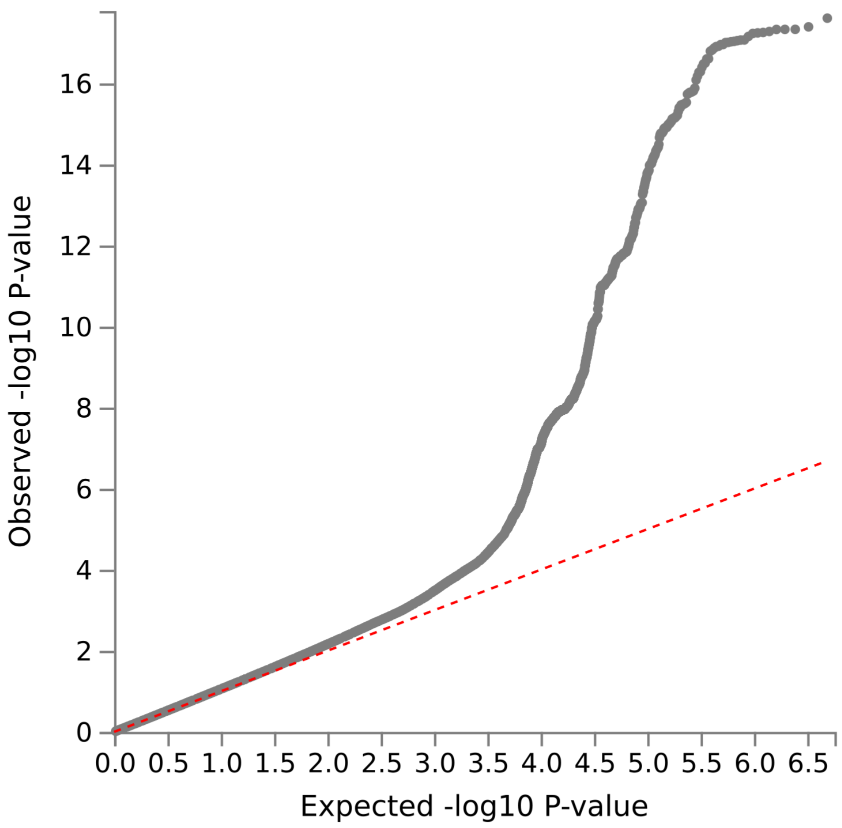


- Lambda GC: 1.0957
- Mean Chi$^2$: 1.117
- Intercept: 1.0238 (0.0069)
- Attenuation ratio: 0.2038 (0.0587)

**Supplementary Figure 2.** GWAS meta-analysis results for (a) sectoral pRNFL, (b) sectoral BMO-MRW, and (c) BMO area. Manhattan plot showing the $-\log_{10}$ (p-values) of association tests in the x-axis. Each point represents an SNP plotted by chromosomal position. The horizontal dashed red line indicates the genome-wide significance threshold (p-value = $5 \times 10^{-8}$).

(a) Sectoral pRNFL

| Temporal | Nasal |
|---|---|
| 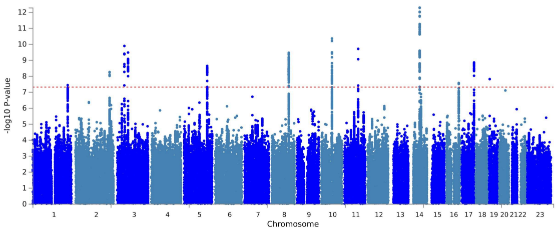  | 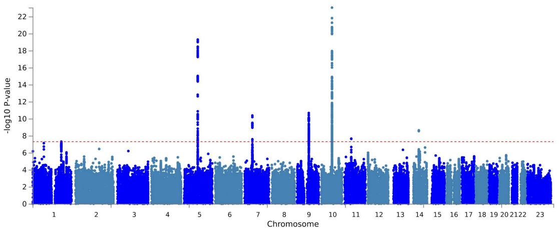  |
| Superotemporal | Superonasal |
| 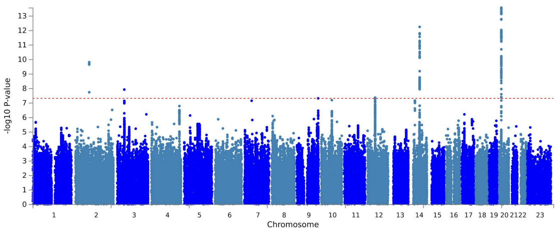  | 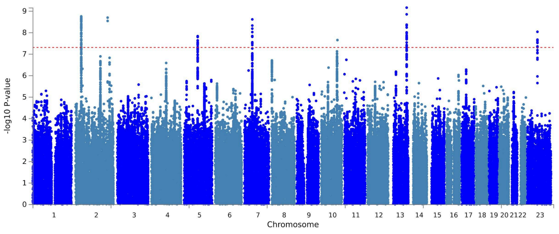  |
| Inferotemporal | Inferonasal |
| 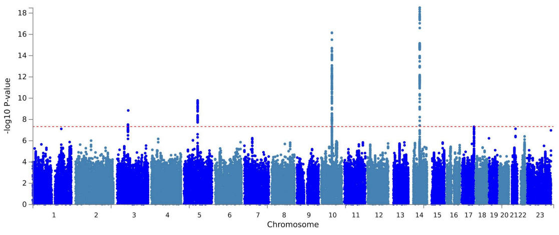  | 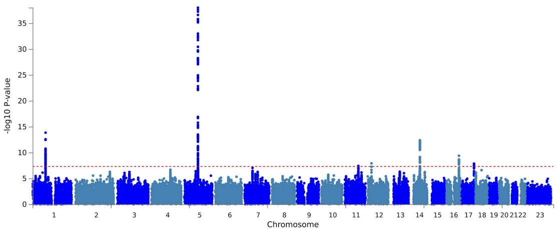  |

(b) Sectoral BMO-MRW

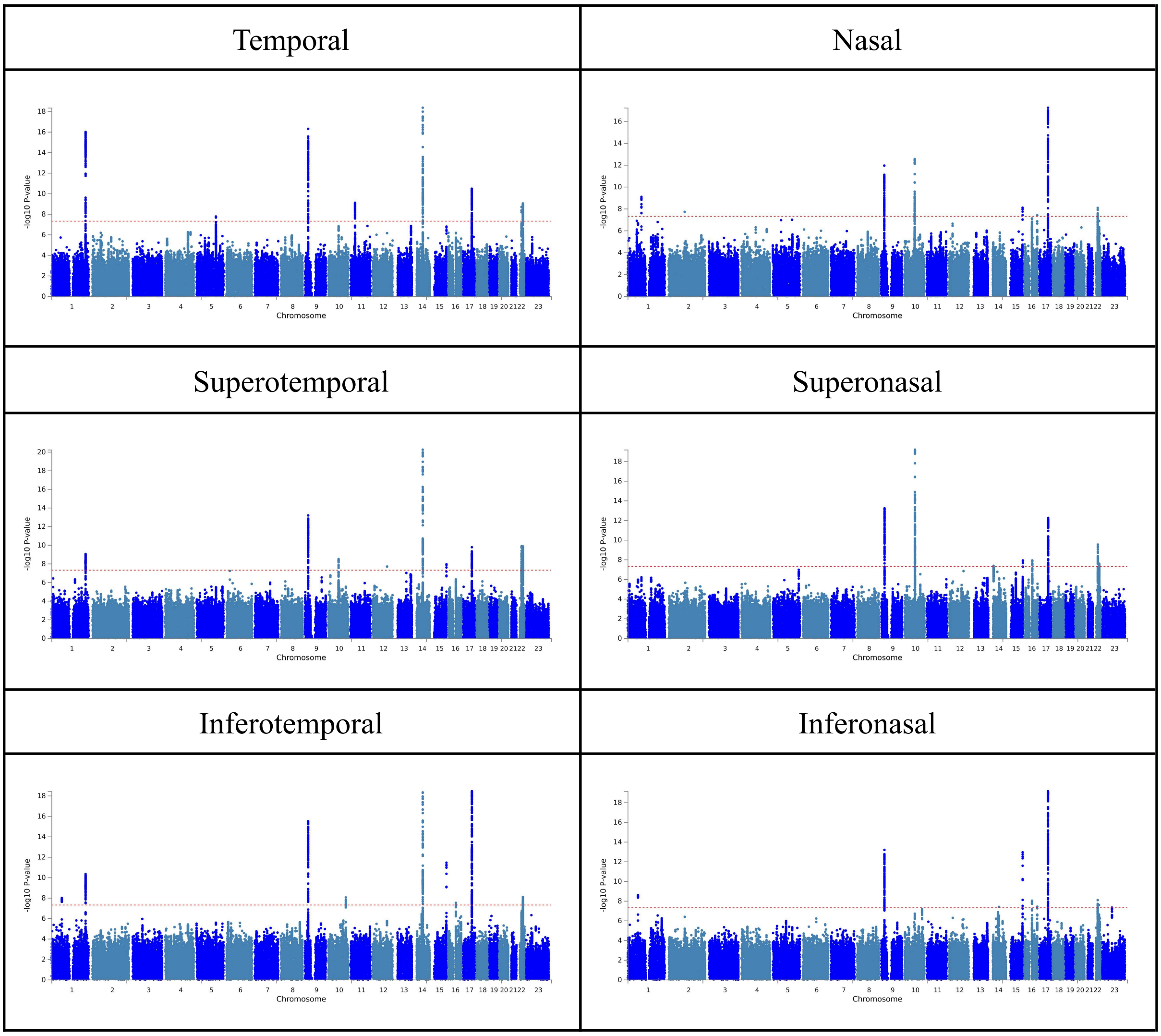


(c) BMO area

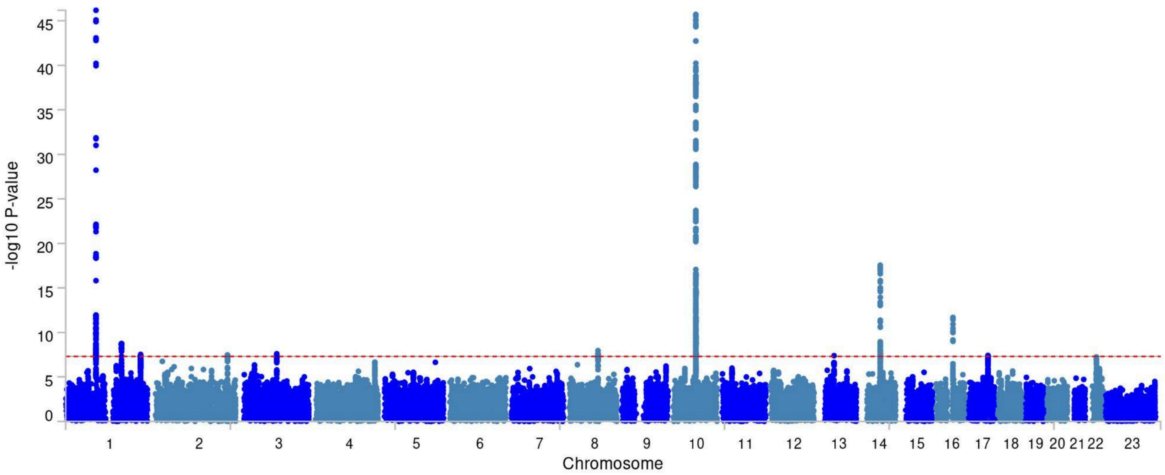

**Supplementary Figure 3.** Conditional analysis results using the mtCOJO method. Manhattan plots provided for (a) IOP-adjusted global pRNFL thickness and (b) IOP-adjusted global BMO-MRW.

(a) IOP-adjusted global pRNFL thickness

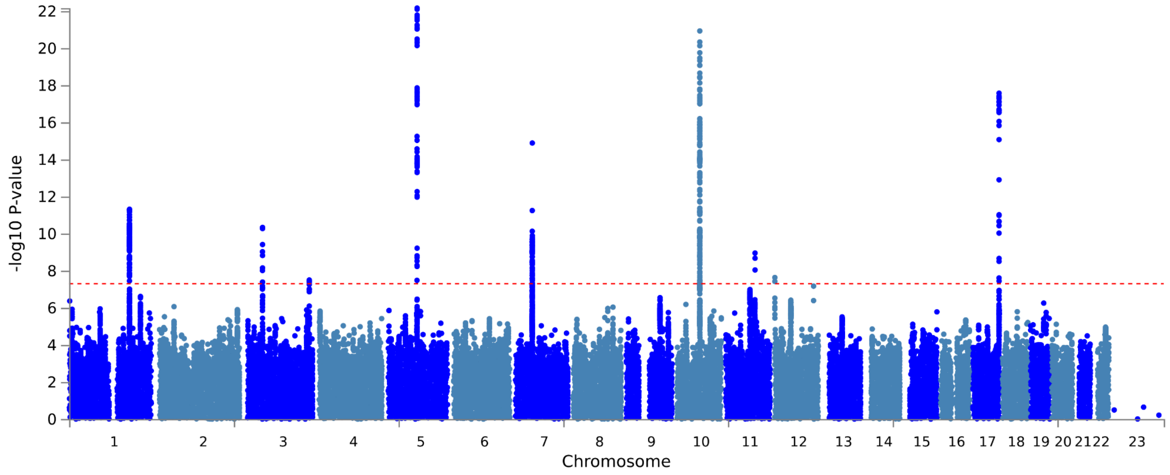


(b) IOP-adjusted global BMO-MRW

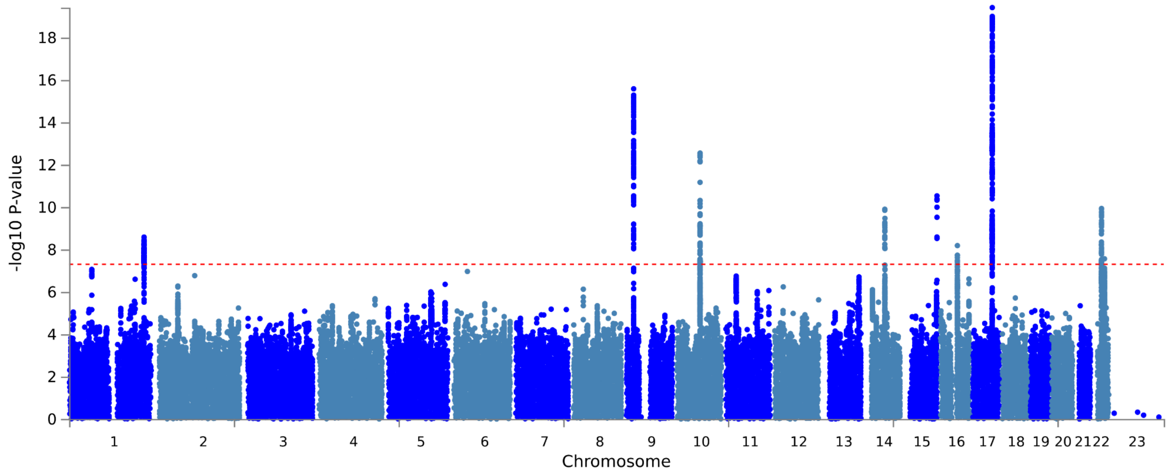

**Supplementary Figure 4.** Mendelian randomisation scatter plots showing the effects of instrumental variables on the exposure (x-axis) and the outcome (y-axis) across different methods.

(a) IOP on global pRNFL thickness

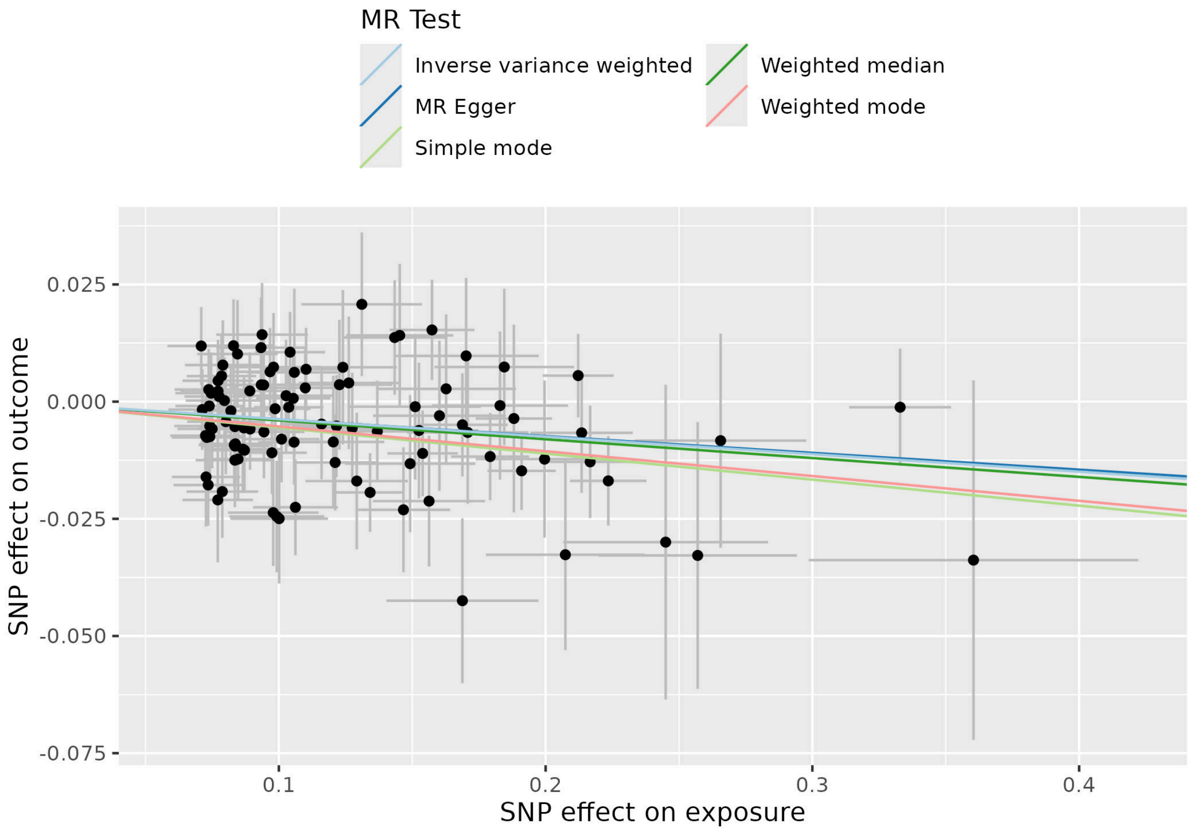

(b) IOP on global BMO-MRW

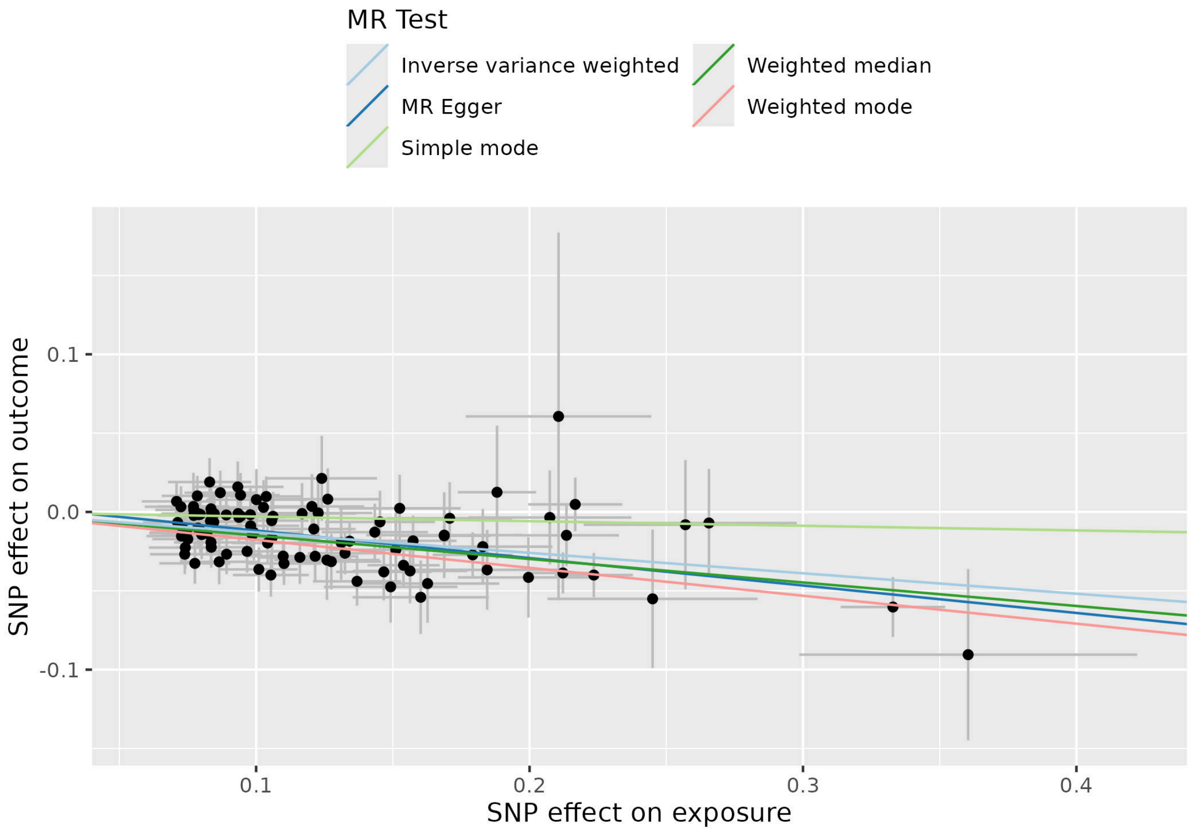
